# Cerebrospinal fluid 2-hydroxyglutarate (2-HG) as a monitoring biomarker for IDH-mutant gliomas

**DOI:** 10.1101/2023.03.01.23286412

**Authors:** Cecile Riviere-Cazaux, Jean M. Lacey, Lucas P. Carlstrom, William J. Laxen, Amanda Munoz-Casabella, Matthew D. Hoplin, Samar Ikram, Abdullah Bin Zubair, Katherine M. Andersen, Arthur E. Warrington, Paul A. Decker, Timothy J. Kaufmann, Jian L. Campian, Jeanette E. Eckel-Passow, Sani H. Kizilbash, Silvia Tortorelli, Terry C. Burns

## Abstract

D-2-hydroxyglutarate (D-2-HG) is a well-established oncometabolite of isocitrate dehydrogenase (IDH) mutant gliomas. While prior studies have demonstrated that D-2-HG is elevated in the cerebrospinal fluid (CSF) of patients with IDH-mutant gliomas^1,2^, no study has determined if CSF D-2-HG can provide a plausible method to evaluate therapeutic response. We are obtaining CSF samples from consenting patients during their disease course via intra-operative collection and Ommaya reservoirs. D-2-HG and D/L-2-HG consistently decreased following tumor resection and throughout chemoradiation in patients monitored longitudinally. Our early experience with this strategy demonstrates the potential for intracranial CSF D-2-HG as a monitoring biomarker for IDH-mutant gliomas.

---

IDH is an important enzyme mutated in most astrocytomas arising in young adults, and all oligodendrogliomas. While IDH should produce alpha-ketoglutarate from the oxidative decarboxylation of isocitrate, the mutant enzyme results in aberrantly elevated reduction of alpha-ketoglutarate to D-2-HG^3^. Overabundant D-2-HG has profound epigenetic effects^4^ and reprograms the glioma metabolome^5^, increasing intracellular oxidative stress. The IDH mutation confers a significant survival benefit to patients with glioma, even in the most aggressive grade 4 astrocytomas (31 months overall survival versus 15 months survival)^6^. As all gliomas inevitably recur, longitudinal monitoring is essential for these patients to enable early detection of tumor recurrence. Unfortunately, surveillance via MR imaging is limited by the frequent presence of pseudoprogression following radiation or immunotherapies and suboptimal sensitivity to early tumor regrowth^7^. Peripheral blood D-2-HG has yielded poor sensitivity to IDH-mutant gliomas^8^. Additionally, although magnetic resonance spectroscopy for 2-HG has shown pharmacodynamic responses to IDH inhibitors^9^, technical challenges have to date impeded clinical application outside of select centers.

CSF is increasingly considered the most robust source for glioma liquid biopsies^10^. Higher 2-HG has been previously reported in CSF of patients with IDH-mutant gliomas than IDH-wild type gliomas, with one study reporting 84% sensitivity and 90% specificity for IDH-mutant gliomas^1^. However, the ability of D-2-HG to serve as a longitudinal monitoring biomarker responsive to therapy remains unknown.

To address this question, patients were enrolled into our Ommaya reservoir study (NCT04692337), which enables longitudinal access to CSF. The catheter for the implanted reservoir was placed within the surgical resection cavity following a standard-of-care resection. Intra-operatively collected CSF samples were also biobanked through Mayo Clinic’s Neuro-Oncology biorepository. If a patient required a ventriculoperitoneal shunt or external ventricular drain, samples were also obtained from these devices through our ongoing CSF biomarkers study (NCT04692324). De-identified patient numbers were assigned to each patient that are unknown to anyone outside of the research team. CSF samples were assayed for D-and-L-2-HG via gas chromatography-mass spectrometry (GC-MS) at Mayo Clinic’s Biochemical Genetics Laboratory. Toward our goal of evaluating 2-HG as a monitoring biomarker for IDH-mutant gliomas, CSF was collected prior to and after resection in six patients (n=5 via Ommaya reservoir and n=1 via external ventricular drain), all of whom underwent near gross total resections (>95% of tumor volume). D-2-HG and the D/L ratio fell on average by 57.9% (range: 15.1%-98.6%) and 68.8% (range: 49.3%-98.9%), respectively, following resection in these six patients (Fig. 1A). The highest percent decrease was observed in a patient with a largely intraventricular grade 4 IDH-mutant astrocytoma (D-2-HG: 99.1%; D/L ratio: 99.3% by POD2), consistent with the impact of ventricular contact on baseline CSF D-2-HG levels (Fig. 1B).

**Figure 1.**
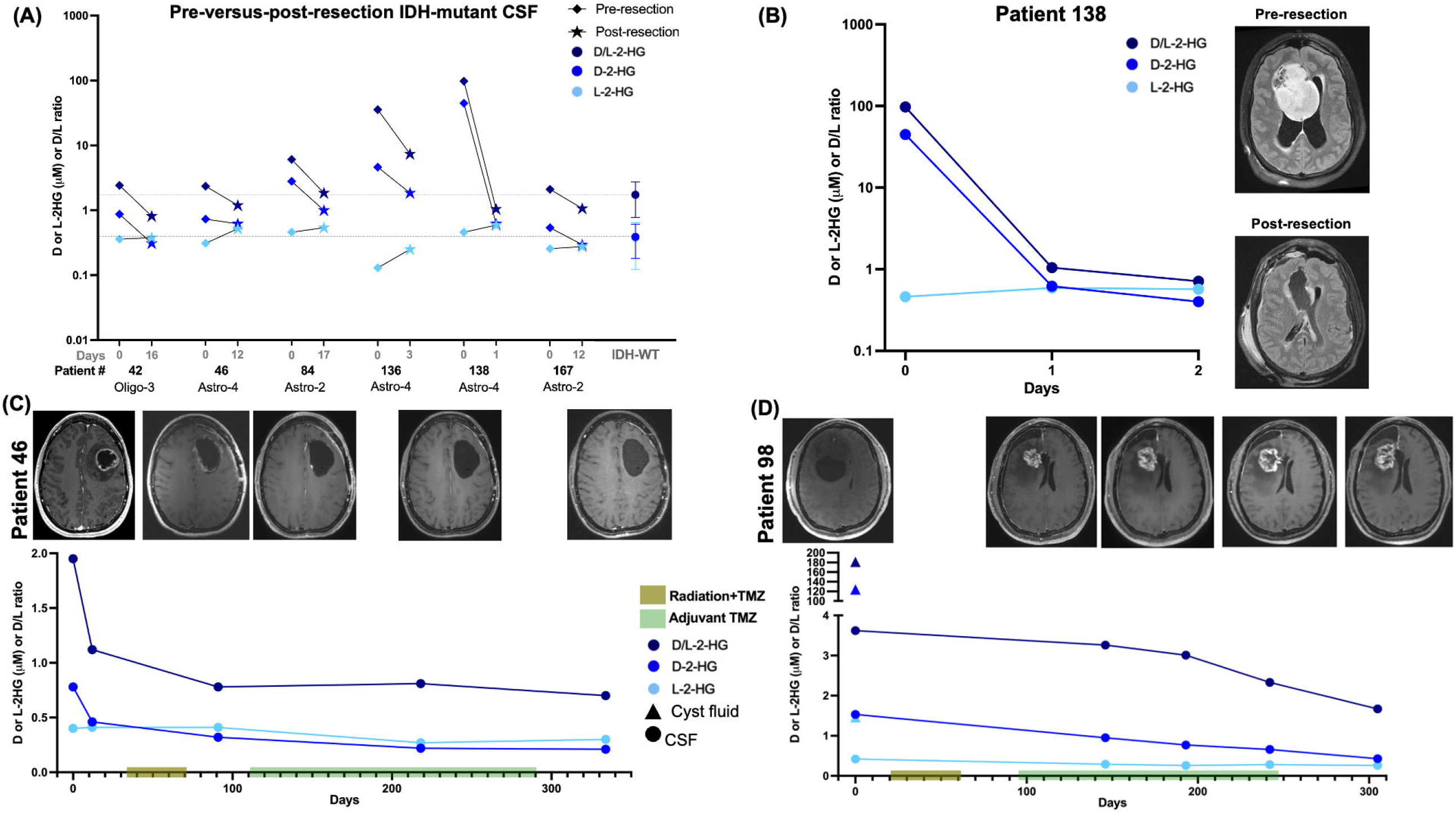
D-2-HG and D/L-2HG decrease with decreasing tumor burden. (A) D-and-L-2-HG were quantified via gas chromatography-mass spectrometry (GC-MS) in cerebrospinal fluid (CSF) samples collected prior to and after resection in seven patients with IDH-mutant gliomas, six of which had primary tumors and one of which had a recurrent glioma (patient 136). The average and standard deviation are shown from 30 intracranial IDH-wild type glioma CSF samples. (B) CSF was obtained from patient 138 (intraventricular grade 4 IDH-mutant astrocytoma) prior to resection and on post-operative days 1 and 2 via an external ventricular drain. CSF was obtained from (C) patient 46 and (D) patient 98 (both grade 4 IDH-mutant astrocytomas) prior to resection and at multiple time points during chemoradiation treatment.

As of the time of this submission, two patients had undergone regular Ommaya reservoir sampling for at least 10 months throughout subsequent chemoradiation. In one patient (Patient 46) undergoing standard of care chemoradiation treatment, D-2-HG and D/L ratio declined slightly during chemoradiation and remained stable throughout adjuvant TMZ, in agreement with stable MR imaging findings (Fig. 1C). In Patient 98, while no post-resection sample was collected prior to their chemoradiation, CSF was collected at the time of surgery and while they were on adjuvant TMZ and an immunotherapy. Despite evidence of presumed pseudoprogression in Patient #98 during treatment on an immunotherapy trial, levels of D-2-HG and the D/L ratio continued to decrease over time (Fig. 1D). These data demonstrate the potential utility of 2-HG monitoring in cases equivocal for pseudoprogression versus progression, particularly for patients who are otherwise clinically stable. Additionally, high levels of D-2-HG and D/L-2-HG were observed in their tumor cyst at the time of resection, consistent with the idea that D-2-HG locally accumulates in fluid spaces directly in contact with tumor. In comparison, CSF obtained at the time of resection from a cortical sulcus distant from the tumor bulk was near the patient’s D-2-HG and D/L-2-HG levels following chemoradiation, suggesting that proximity to the tumor may impact measured levels of these metabolites.

Sample collection is ongoing for all patients. Thankfully, none of our patients have yet experienced a true recurrence of their IDH-mutant glioma. As such, we have not yet been able to independently confirm recent findings of increasing CSF D-2-HG with tumor recurrence in two patients^2^. Nevertheless, our data provide the most complete evidence to date regarding the potential dynamic range of D-2-HG achievable within individual patients for longitudinal monitoring and response assessment.

In conclusion, we have demonstrated proof-of-principle data from our initial patient cohort regarding the ability of CSF D-2-HG to reflect decreased tumor burden. Work is ongoing to evaluate the reproducibility of these findings in a larger cohort of patients with low or high-grade gliomas. Future studies will evaluate the impact of IDH inhibitors and other experimental therapies on CSF 2-HG levels for pharmacodynamic monitoring of treatment response. Ultimately, we expect that CSF 2-HG will provide early insights into treatment response and disease progression in patients with IDH-mutant gliomas, particularly when combined with other biomarkers of cytotoxicity and treatment response. It is worth noting that even with intracranial monitoring, D-2-HG levels approximated median background levels after surgery in 2/7 patients, even prior to chemoradiation. Cranial CSF D-2-HG levels were previously reported to exceed those obtained in lumbar CSF. As such, intracranial CSF may provide maximal sensitivity for longitudinal monitoring of IDH-mutant gliomas via D-2-HG.

## Data Availability

All data produced in the present study are available upon request to the author.

## Notes

**Conflict of Interest:** None.

### Competing Interest Statement

The authors have declared no competing interest.

### Funding Statement

This study was funded by: National Institutes of Health NINDS R61 NS122096 to TCB, TJK, JEP, PAD, SHK, AEW; National Institute of Health T32GM065841 to CRC.

### Author Declarations

The Mayo Clinic Institutional Review Board at Rochester, MN approved this study.

